# The reliability of two prospective cortical biomarkers for pain: EEG peak alpha frequency and TMS corticomotor excitability

**DOI:** 10.1101/2022.03.06.22271797

**Authors:** Nahian S Chowdhury, Patrick Skippen, E Si, Alan Chiang, Samantha K Millard, Andrew J Furman, Shuo Chen, David A Seminowicz, Siobhan M Schabrun

## Abstract

Many pain biomarkers fail to move from discovery to clinical application, attributed to poor reliability and feasible classifications of at-risk individuals. Preliminary evidence has shown that higher pain sensitivity is associated with slow peak alpha frequency (PAF) and depression of corticomotor excitability (CME). The present study evaluated the reliability of these measures, specifically whether, over several days of pain, a) PAF remains stable and b) individuals show two stable and distinct CME responses: facilitation and depression. Seventy-five healthy participants were given an injection of nerve growth factor (NGF) into the right masseter muscle on Day 0 and Day 2, inducing sustained pain. Electroencephalography (EEG) to assess PAF and transcranial magnetic stimulation (TMS) to assess CME were recorded on Day 0, Day 2 and Day 5. PAF reliability was in the excellent range even without standard pre-processing and ∼2 minutes recording length. Moreover, two distinct and stable CME responses were demonstrated: facilitation and depression. These findings support the notion that PAF is a stable trait characteristic, with reliability unaffected by pain, and excellent reliability achievable with minimal pre-processing and ∼2 minutes recording, making it a readily translatable biomarker. Furthermore, the study showed novel evidence of two stable corticomotor adaptations to sustained pain. Overall, the study provides support for the reliability of PAF and CME as prospective cortical biomarkers.

---

Chronic pain continues to be a major source of disability worldwide. As a result of the significant challenges associated with current pain management approaches, many researchers have shifted focus to the development of biomarkers that can be used in the diagnosis, prevention and treatment of chronic pain [1, 2]. However, many prospective pain biomarkers fail to move from the initial discovery stage to clinical application [1]. One reason for this failure is poor biomarker reliability and feasible classifications of identify at-risk individuals [1, 3]. Initial evaluations of biomarker reliability and feasibility are vital, as they determine the feasibility of subsequent stages of biomarker development, including clinical validation and intervention studies.

Recent work has shown promise for a cortical biomarker pain signature involving two measures: peak alpha frequency (PAF), which is measured using resting-state electroencephalography (EEG) and refers to the dominant oscillatory frequency in the 8-12Hz range [4-6] and; corticomotor excitability (CME), which is measured using transcranial magnetic stimulation (TMS), and can be indexed as the volume of the primary motor cortex (M1) representation of a peripheral muscle [7]. Studies using pain models in healthy individuals have shown that PAF and CME can predict pain severity and duration [8-12]. One of these studies used intramuscular injections of Nerve Growth Factor (NGF), which produces progressively developing and clinically meaningful pain sustained over several days. Within the same sample, it was shown that those with slower PAF (lower frequency) prior to pain, and reduced CME during pain (“depressors”) experienced higher pain and disability, while those with faster PAF (higher frequency) and increased CME (“facilitators”) experienced less pain and faster recovery [13]. These preliminary findings suggest that the combination of reduced CME and slow PAF could serve as a biomarker that can identify individuals at risk of developing longer lasting and more severe pain following an acute injury.

Though CME and PAF hold promise as biomarkers, there are outstanding questions regarding their reliability. One question pertains to the reliability of PAF over several days of sustained pain. Experimental pain has been shown to influence PAF and alpha power [14-16]. As such, the presence of pain may introduce fluctuations in PAF that could affect the usefulness of pre-pain classification methods. While Furman and colleagues [8] demonstrated excellent reliability of sensorimotor PAF over several days of sustained pain, an in-depth investigation of the methodological factors that may influence PAF reliability was not conducted, including the pre-processing pipeline used to reduce EEG signal noise [17, 18], the frequency window used to calculate PAF [11], the duration of the EEG recording [19], how the peak frequency is identified [5], and the sensors/region of interest chosen to calculate PAF [20]. Such an investigation would ascertain whether the reliability of PAF during pain remains similar regardless of these methodological considerations or whether reliability is compromised under certain conditions.

Another outstanding question is whether individuals reliably show two distinct patterns of CME change during pain: corticomotor facilitation and depression. Many theories of chronic pain suggest that individuals show significant variations in motor adaptations in response to pain sustained over long periods, with some of these adaptations associated with increased risk of chronic pain [21, 22]. As it is difficult to assess chronic pain patients during the transition period, the use of experimental pain models that produce long lasting and clinically relevant pain in healthy participants provide a convenient method to identify these individual motor strategies. However, only one study has characterised the individual level responses in CME during pain sustained over several days [9], which showed that facilitators represented ∼40% of participants, while depressors represented 60%. Moreover, this study focused on upper limb pain, whereas corticomotor adaptations in other body regions may differ. One such region is the masseter muscle, which in contrast to the upper limb, is involved in different motor functions and is subserved by trigeminal motoneurons rather than spinal motoneurons [23, 24]. Demonstrating two distinct responses to pain (facilitation and depression) in the masseter muscle would improve the generalizability of CME classifications.

The aim of the present study was to a) evaluate the reliability of PAF over several days of sustained pain and b) determine whether individuals show two stable and distinct patterns of CME change during sustained pain: corticomotor facilitation and depression. To address these aims, we used a subsample of participants (n = 85) from PREDICT, an ongoing initial biomarker validation study for CME and PAF [25]. Participants were administered with an intramuscular injection of NGF to the right masseter muscle at baseline (Day 0), and two days after baseline (Day 2). CME and PAF were measured on Day 0 (prior to the injection), Day 2 and Day 5.

## Methods

### Design

This paper was an interim analysis of the PREDICT trial [25] that aims to undertake analytical validation of the PAF and CME biomarkers in a human model of temporomandibular disorder (TMD). A longitudinal experimental design was used, where participants experienced the development and complete resolution of NGF-induced right masseter muscle pain, with outcomes collected over a period of 30 days. The PREDICT trial is prospectively registered on ClinicalTrials.gov (NCT04241562) and a protocol paper has been published [25].

### Participants

As the aim of the PREDICT study was to mimic the symptoms of temporomandibular disorder, the inclusion criteria were based on the OPPERA prospective cohort study [26], which demonstrated first-onset TMD incidence rate of 2.5%-4.5% per annum among 18 to 44-year-olds, and a marginal difference in TMD incidence between males and females. As such, this study included the first 85 participants who completed the study (38 females, 47 males; age range 18-43, mean 25.72 ± 5.96). Participants were excluded for the following reasons: 1) inability or refusal to provide written consent, 2) presence of acute pain, 3) history or presence of acute or chronic pain, 4) history or presence of a medical or psychiatric condition, 5) frequent use of alcohol, opioids, or illicit drugs in the past 3 months, 6) pregnant or lactating women, 7) contraindicated for TMS (e.g., metal implants, epilepsy). Full details are included in the published protocol [25]. Participants were recruited via advertisements placed on community notice boards, social media platforms and a healthy participant volunteer database. Ethical approval was obtained from the University of New South Wales (HC190206) and the University of Maryland Baltimore (HP-00085371). All procedures were conducted in accordance with the Declaration of Helsinki. Written, informed consent was obtained, and participants were free to withdraw from the study at any time.

### Experimental Protocol

Participants attended the laboratory on Day 0, Day 2, and Day 5, with facial pain, PAF and CME measured at each session. NGF was injected into the right masseter muscle at the end of the Day 0 and Day 2 laboratory sessions. As part of the larger PREDICT study, other data (not reported here), included electronic pain diaries collected from Day 0 to Day 30, questionnaire data collected on Day 0, Day 2 and Day 5, and pressure pain thresholds obtained on Day 0, Day 2 and Day 5.

### Data collection procedures

#### Facial Pain Questionnaires

At the beginning of each session, participants rated their facial pain on an 11-point numerical rating scale at rest, chewing, swallowing, drinking, talking, yawning and smiling. The scale ranged from 0 = “no pain” to 10 = “worst pain imaginable”.

#### Peak alpha frequency (PAF)

Participants were seated in a comfortable chair. Scalp EEG was recorded using the Brain Vision Recorder (ActiChamp Plus, Brain Products GmbH, Germany). Recording was done at a sampling rate of 25000 Hz (participants 1-16) or 5000 Hz (participants 17-85). Signals were recorded from 63 active wet electrodes, embedded in an elastic cap (ActiCap, Brain Products GmbH, Germany) in line with the 10-10 system. Recordings were referenced online to ‘FCz’ and the ground electrode placed on ‘FPz’. Electrode impedances were maintained below 25 kOhms during setup. Once setup was complete, the lights were switched off with ambient noise reduced to a minimum. Participants were instructed to relax their muscles and keep their eyes closed while remaining awake. The resting-state EEG signal was then recorded for 5 minutes.

#### Corticomotor excitability (CME)

##### Electromyography

Participants were seated in a comfortable chair and viewed a monitor that displayed electromyographic (EMG) feedback. Bipolar surface electrodes were used to record EMG from the right masseter muscle. A belly-tendon montage was used, with the active (belly) and reference (tendon) electrodes placed along the mandibular angle, and ground electrode placed on the right acromion. EMG signals were amplified (x 1000) and filtered (16 to 1000Hz), and digitally sampled at 2000 Hz.

##### Maximum Voluntary Contraction

It is common to assess CME from the masseter muscle during active contraction that is at a certain percentage of the maximum voluntary contraction (MVC) (see [23, 24]). To measure MVC, participants were actively encouraged to clench their jaw as hard as possible for 3 seconds on 3 separate trials, with a 1-minute rest break in between trials. The average and 20% MVC were computed. Participants were provided with practise at maintaining a masseter contraction of 20% MVC by receiving live feedback from the EMG signal.

##### Transcranial Magnetic Stimulation Hotspot

Single, monophasic stimuli were delivered using a Magstim unit (Magstim Ltd., UK) and 70mm figure-of-eight flat coil. In line with previous studies investigating optimal coil orientation for inducing masseter MEPs, an angle of 90 degrees between the anterior-posterior line and the coil handle was used [27]. This orientation induced a current in the lateral-to-medial direction. Participants wore a swim cap with a grid of 1cm x 1cm resolution. The scalp site evoking the largest MEP (i.e., the “hotspot”) during active contraction was then determined.

##### Thresholding

The TMS motor threshold assessment tool [28] was used to determine the active motor threshold (AMT), defined as the minimum intensity required to evoke a reliable masseter MEP during active contraction. A reliable MEP was identified if the EMG waveform between ∼5-15ms [29, 30] after the TMS pulse was visibly larger in amplitude relative to background EMG.

##### Mapping

The procedure for mapping has been described in detail previously [31, 32]. The TMS intensity was set at 120% AMT. During active contraction, 3 stimuli were delivered at each location around the grid, starting at the hotspot. The number of stimulation sites was pseudorandomly increased until an MEP was no longer observed (i.e., no reliable MEP in all 3 trials at all border sites). Note that on Day 2 and 5, the mapping procedure began immediately after the MVC was determined, with the motor hotspot and threshold procedures skipped. The same hotspot and test stimulus intensity as determined on Day 0 was used for the mapping procedures on Day 2 and Day 5.

#### Intramuscular injection of nerve growth factor (NGF)

The NGF injections were provided at the end of the Day 0 and Day 2 sessions. To prepare the masseter muscle for injection, the surface was cleaned using alcohol wipes. A sterile solution of recombinant human NGF (dose of 5 μg [0.2 ml]) was then administered as a bolus injection into the muscle belly of the right masseter using a 1-ml syringe with a disposable needle (27-G). The needle was inserted perpendicular to the masseter body until bony contact was reached, retracted ∼2mm, and NGF injected. To confirm that this injection induced sustained pain over several days, we obtained participants pain ratings (numerical rating scale score /10) at rest, and upon chewing, for the Day 2 and 5 sessions.

### Data processing

#### Peak Alpha Frequency

EEG data processing was conducted using custom MATLAB (R2020b, The Mathworks, USA) scripts implementing the EEGLAB (eeglab2019_1) [33] and Fieldtrip (v20200215) toolboxes [34]. We investigated the influence of four factors on the reliability of PAF at each channel: pre-processing pipeline, recording length analysed, choice of frequency window, and calculation method.

##### Impact of Pipeline

Two pre-processing pipelines were compared. These pipelines were labelled the “Standard cleaning” and “No cleaning” pipelines. The standard cleaning pipeline matched those used in previous studies investigating the relationship between PAF and pain: data downsampled to 500Hz, re-referenced to average, and band-pass filtered between 2 and 100Hz using an FIR filter. Channel data was visibly inspected, and overtly noisy channels were removed, and the signal re-referenced. Data were segmented into 5 second epochs, and epochs containing marked muscular artefacts were visually rejected. Principal component analysis was applied to identify and remove components relating to eye blinks and/or saccades. Removed channels were then interpolated using the nearest neighbour method [34]. In the “no cleaning” pipeline, the data was downsampled to 500 Hz, re-referenced to average, and segmented into 5 second epochs. For both pipelines, the power spectral density was derived in .2Hz bins. The 2-50Hz range was extracted using Fast Fourier Transform. A Hanning taper was applied to the data to reduce edge artefacts.

##### Impact of Recording Length

Two recording lengths were analysed: the first half of the remaining epochs vs. all the remaining epochs after excluding bad trials.

##### Impact of Frequency Window

The peak frequency in the alpha range was calculated across two frequency windows: a wider 8-12 Hz window vs. a narrower 9-11 Hz window.

##### Impact of Peak Identification Method

Two methods of calculating PAF were compared. One was the peak picking method [35], which is the single frequency that yielded the largest power within the 8-12 or 9-11 Hz range. The other was a weighted frequency approach, known as the Centre of Gravity method, calculated using the following equation:

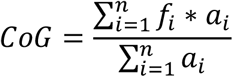

Where f_i_ is the ith frequency bin within the frequency window of interest, and a_i_ represents the spectral amplitude at f_i_.

Test-retest reliability was assessed by computing the intraclass correlation coefficient (ICC, two-way fixed, single measure) [36] of PAF at each electrode across the three sessions. An ICC ≤ 0.2 indicates poor relative reliability, 0.21 to 0.4 fair, 0.41 to 0.6 moderate, 0.61 to 0.8 good, and ≥ 0.81 excellent relative reliability [37].

#### Corticomotor excitability

TMS data was processed using a custom MATLAB script. For 15 randomly selected participants, the onsets and offsets of the MEPs were manually determined for each trial. The mean onset and offset across these participants were then used to fix the MEP onsets and offsets across all participants. To confirm if this was reliable, we determined the correlations in the key outcome measure between the fixed MEP window method vs. the manual selection method. The root mean square (RMS) for each MEP window was determined. The RMS of background EMG was determined using a fixed window between 55 and 5ms before the TMS pulse [38]. This was subtracted from the RMS of the MEP window to determine MEP amplitude. The mean MEP amplitude at each stimulation site was determined. Map volume was calculated by summing the amplitudes of sites exhibiting >10% of the maximum MEP amplitude [39].

For each participant, a CME change score was determined by computing the map volume for Day 2 and Day 5 as a proportion of Day 0 [9]. To determine the reliability of these change scores, the ICC between the Day 2 and Day 5 change score was computed.

To determine whether there were two distinct CME responses, the proportion of participants that showed a meaningful increase (facilitator) or decrease (depressor) in CME was determined. This was based off the mean change score across Days 2 and 5, in line with previous research [9]. Using the sample size and standard deviation in the mean change scores, a “cut-off” score was determined based on a one-sample t-test. A chi-squared test was conducted to determine whether a significant proportion of participants demonstrated an increase or decrease in map volume beyond the cut-off score.

## Results

### Pain Characteristics

On Day 0, all participants reported no pain in the right cheek muscle at rest or chewing. On Day 2, the mean pain score (numerical rating scale score/10) was 1.13 ± 1.54 for pain at rest, and 2.47 ± 2.26 for pain upon chewing. On Day 5, the mean pain score was 0.93 ± 1.26 for pain at rest, and 2.61 ± 1.73 for pain upon chewing.

### Reliability of Peak Alpha Frequency

Ten participants had missing EEG data on Day 0, 2 or 5 due to illness or unwillingness to continue the procedures. This left 75 participants in the final analysis on PAF. For the standard cleaning pipeline, the mean number of channels excluded per recording was 0.5 (SD = 1.09, range = 0-8). The mean number of bad epochs excluded per recording was 1.26 (SD = 2.44, range = 0-18). Given the range of remaining epochs, this suggests that for the standard cleaning pipeline, the remaining full and half recording lengths were on average, close to 5 and 2.5 minutes respectively, with a range of 3.5-5 and 1.75-2.5 minutes respectively. The mean number of eye movement components removed per recording was 1.41 (SD = 0.75, range = 0-2).

Figure 1 shows the individual and group level z-transformed spectral plots (averaged across electrodes) for each day, and for both standard and non-standard cleaning pipelines.

**Figure 1.**
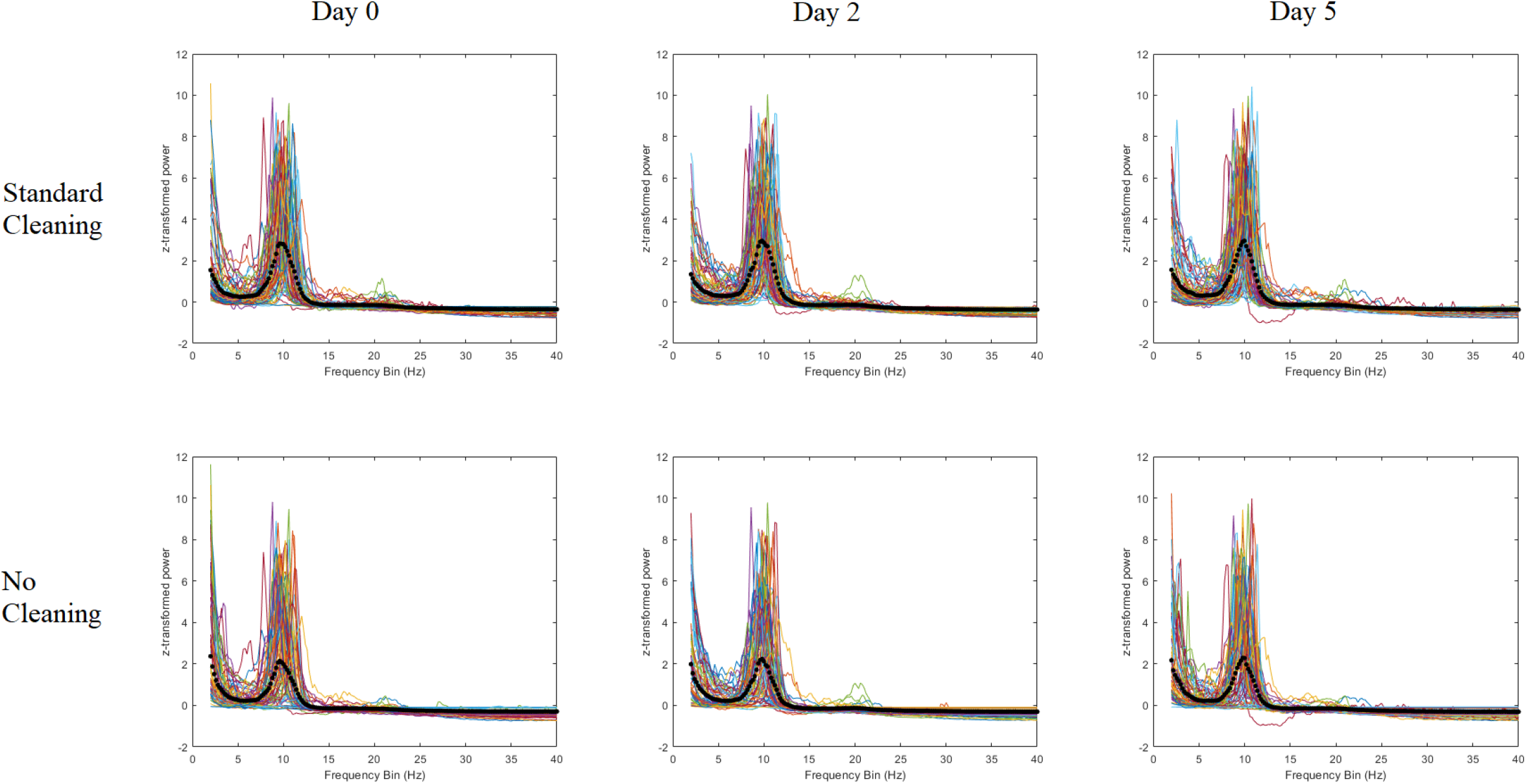
Individual and mean (n=75, in black) EEG spectral plots (averaged across electrodes) for each day, and for both standard and non-standard pre-processing pipelines.

Figure 2 shows topographies and scatterplots for ICCs in PAF as a function of pipeline, recording length, frequency window and calculation method. When using the Centre of Gravity Method, ICCs were in the “excellent” range for almost all channels regardless of frequency window, pipeline, or recording length. For the peak picking method, ICCs were mostly in the “good” range, though the occurrence of “moderate” range ICCs was higher when using an 8-12Hz compared to the 9-11Hz frequency window. These findings suggest that while the use of the Centre of Gravity method leads to excellent reliability regardless of methodological considerations, reliability for the peak picking method varies between moderate to good depending on whether a wider or narrower frequency window is used to calculate PAF.

**Figure 2.**
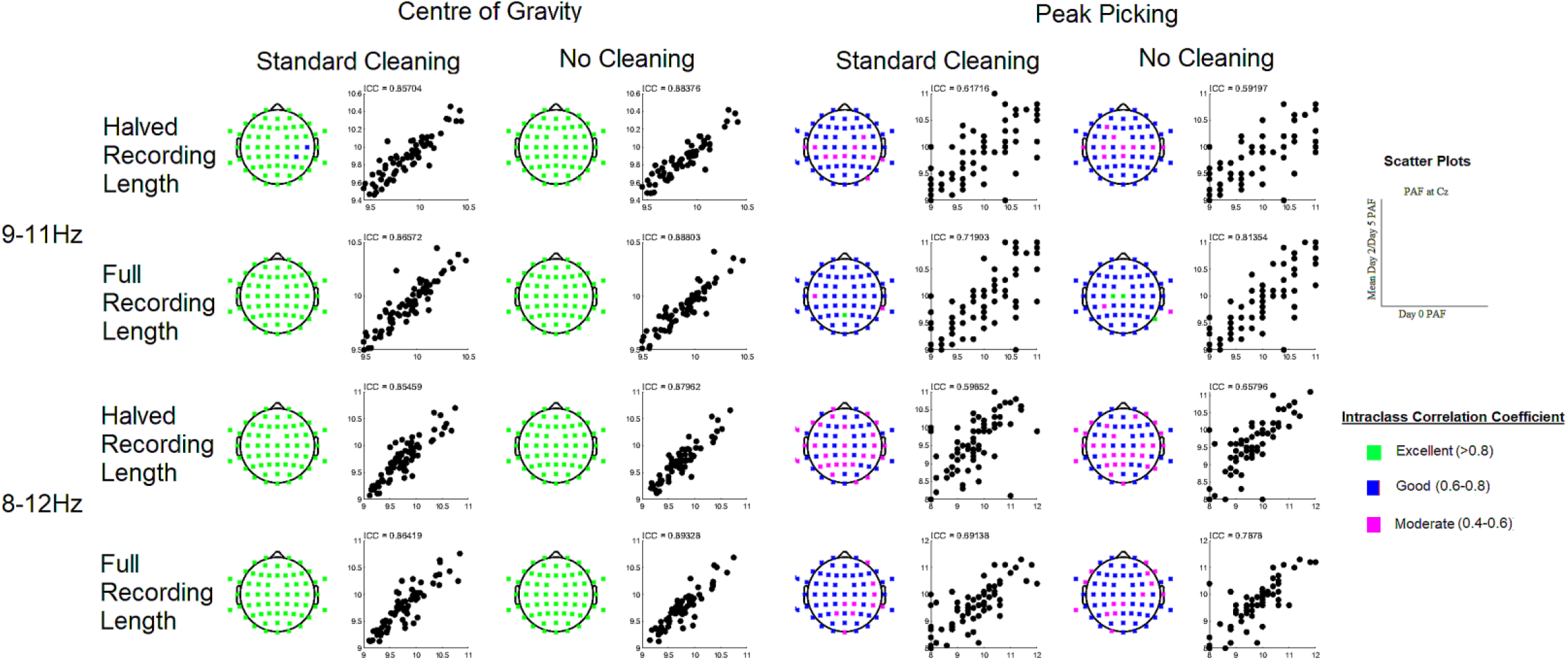
Topographies and scatterplots for ICCs in PAF as a function of pre-processing pipeline, recording length, frequency window and calculation method. Respectively, green, blue, magenta and red represent excellent, good, moderate or poor test-retest reliability of PAF between sessions. The scatterplots show PAF on Day 0 plotted against the mean PAF of Day 2 and Day 5 for a representative channel (Cz). PAF calculated with the COG method had excellent reliability, regardless of the pre-processing pipeline or recording length.

### Changes in Corticomotor Excitability

Eleven participants had missing TMS data on Day 0, 2 or 5 due to illness or unwillingness to continue the procedures. This left 74 participants in the final analysis on CME. The mean MVC and 20% MVC values were 0.048 V (±0.036) and 0.009 V (0.007), respectively. The mean AMT and test stimulus intensities were 42.78 (8.50) and 51.38 (10.18), respectively.

Figure 3 shows an exemplar masseter MEP. MEP windows were selected manually for 15 participants (45 sessions in total).

**Figure 3.**
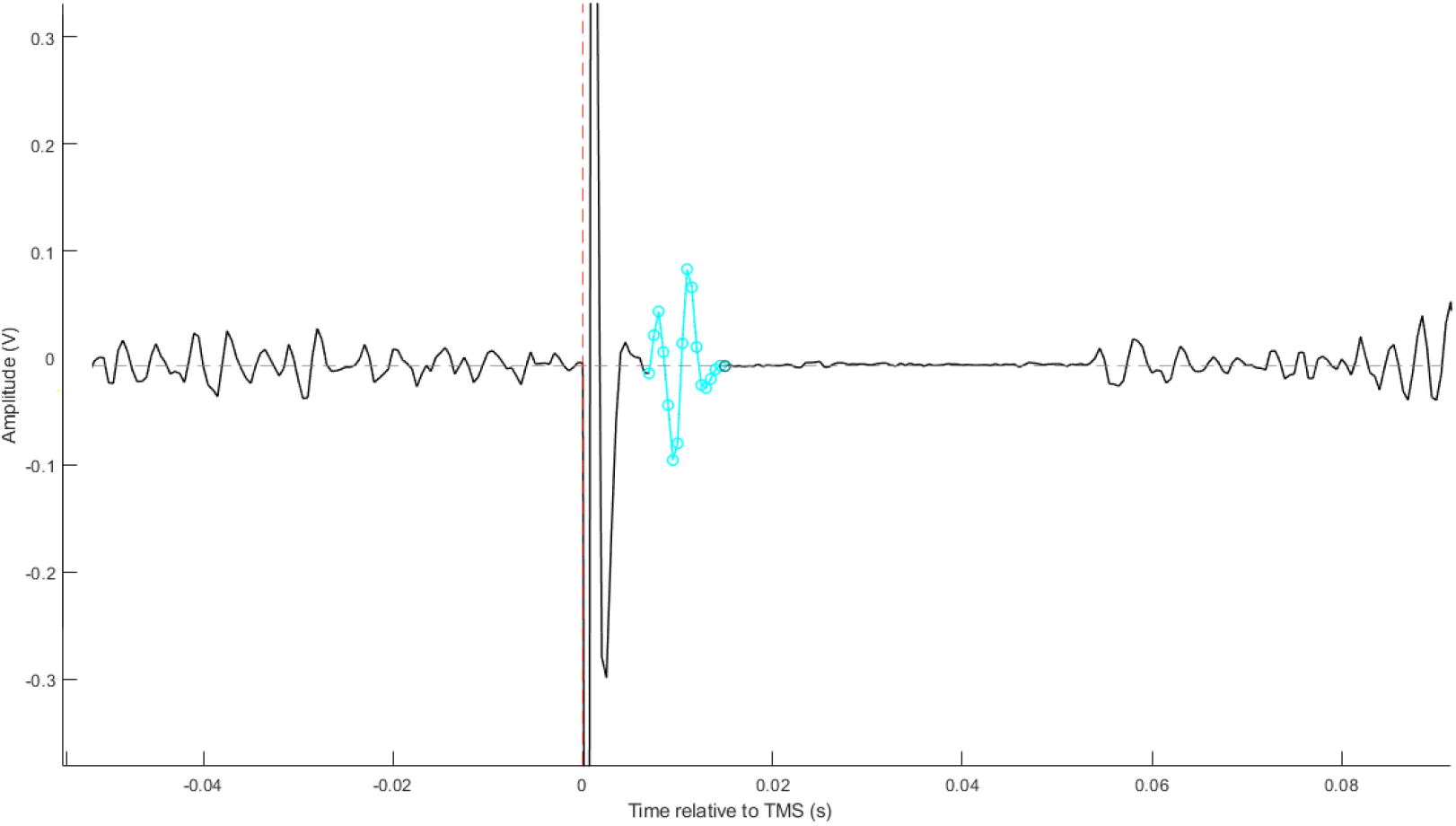
An exemplar masseter MEP. The window between 5 and 55ms before the TMS pulse represents the background EMG window. The blue highlighted window shows the window between the onset and offset of the MEP.

Figure 4 shows a scatterplot of map volumes when MEP windows were selected manually vs. fixed windows. The R^2^ of .99 suggests both methods led to near-identical estimates of map volume, supporting the reliability of the fixed window method.

**Figure 4.**
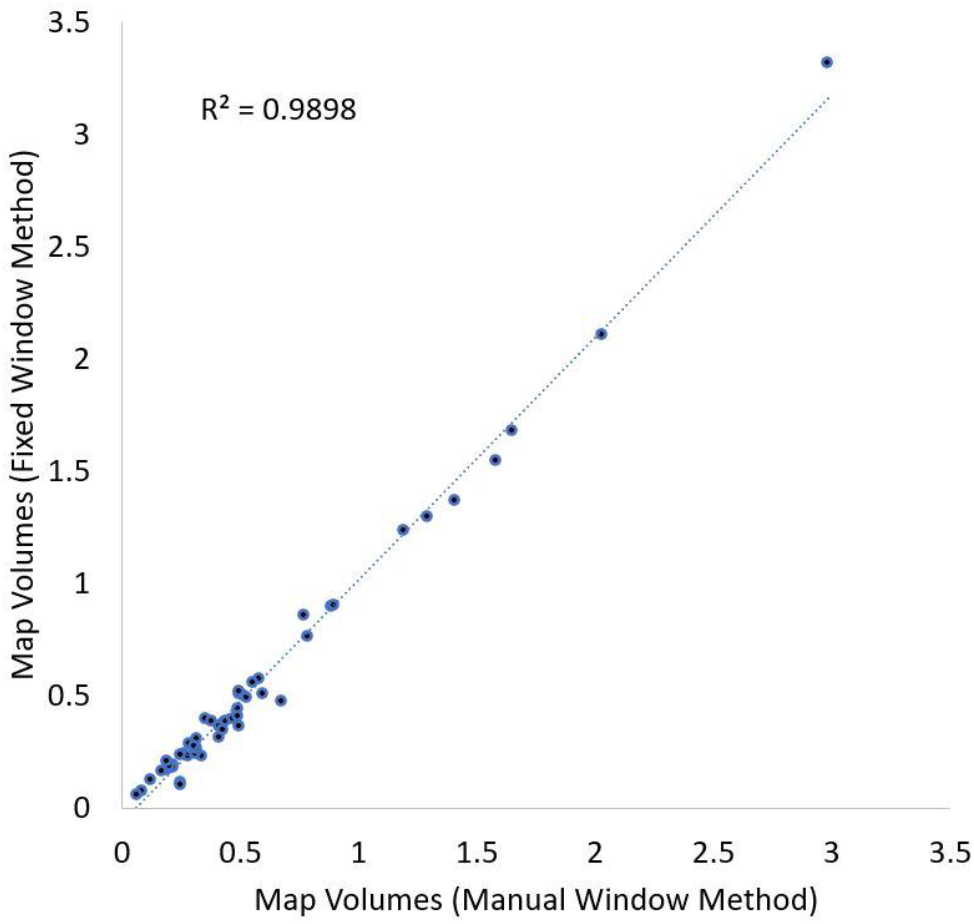
A scatterplot of 45 map volume estimates when MEP windows were selected manually vs. fixed windows.

Shapiro-Wilk tests showed that scores on map volume change (as a proportion of Day 0) violated assumptions of normality for both Day 2 (*W* = 0.66, *p* < .001) and Day 5 (*W* = 0.77, *p* < .001). As such, these change scores were log-transformed, after which assumptions of normality were satisfied for Day 2 (*W* = 0.98, *p* = .29) and Day 5 (*W* = 0.99, *p* = 0.69). Figure 5 plots the log-transformed change scores on Day 2 against Day 5. The ICC between these measures was 0.63, suggesting the stability of the CME response to pain was in the good range.

**Figure 5.**
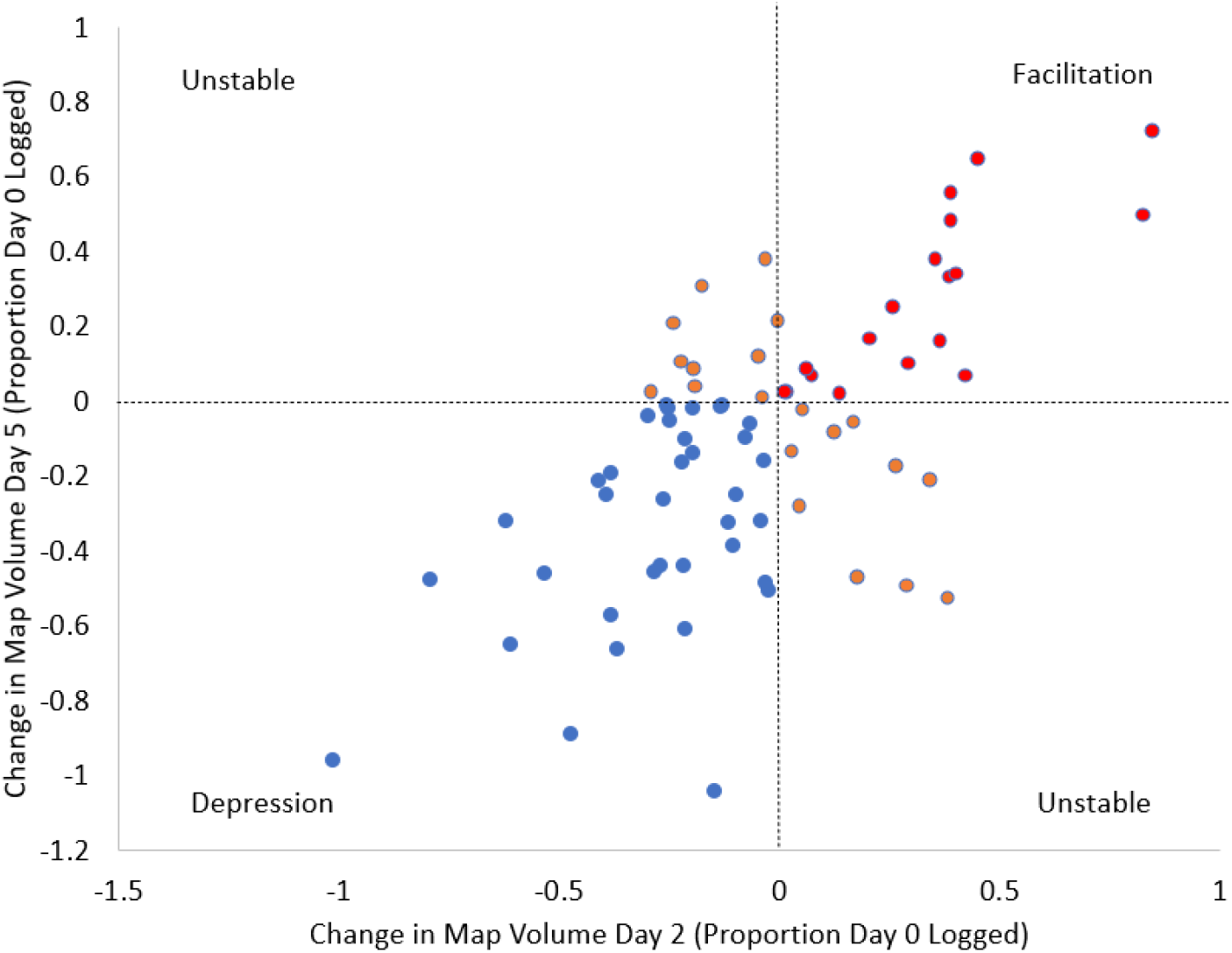
Corticomotor map volume change scores on Day 2 plotted against change scores for Day 5. The blue plots show participants who demonstrated a reduction in map volume on both Day 2 and 5, while the red plots show the opposite pattern. The orange plots show participants that demonstrate “unstable” changes i.e., a decrease on one day but an increase on the other.

Figure 6 shows the change scores for each participant on Day 2 and Day 5 as a proportion of Day 0, with red lines representing facilitators (those who show a mean increase in map volume) and blue lines representing depressors (those who show a mean decrease in map volume).

**Figure 6.**
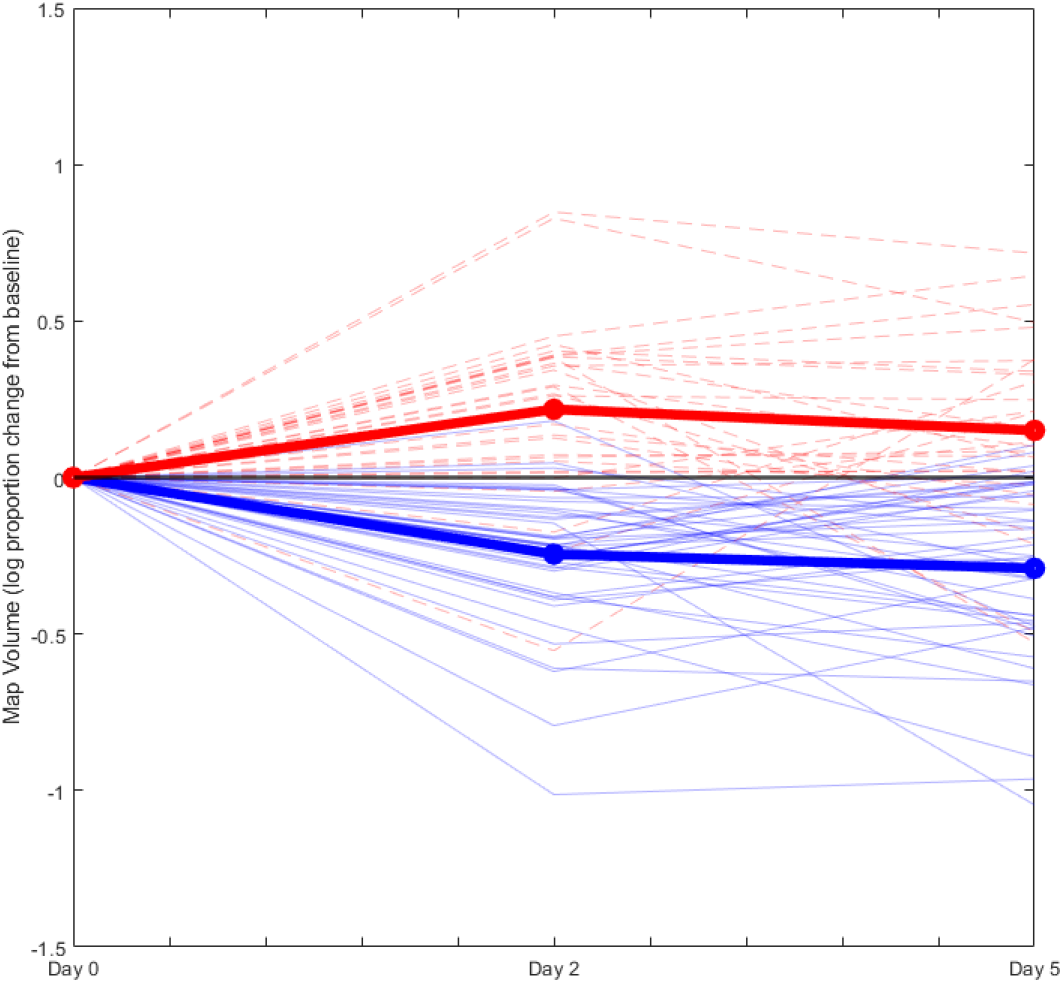
CME change scores for each participant on Day 2 and Day 5 as a proportion of Day Respectively, the light red and blue lines indicate those who showed a mean increase or decrease (across both days) in map volume, and the dark red and blue lines shows the average of each of these groups.

Figure 7 shows the motor cortex maps for each group. These maps display the mean unadjusted MEP amplitudes at each stimulated location. Using the sample size and standard deviation in the mean change score (mean of Day 2 and Day 5 as a proportion of day 0), a “cut-off” was determined that represented a meaningful change in map volume from Day 0 based on a one-sample t-test. This cut-off score was ± 0.0715 on the log scale, which when converted back to the linear scale, corresponded to ∼17% change in map volume relative to day 0. The proportion of individuals demonstrating a change beyond the cut-off score was 58/74, which represented a significant proportion of individuals (χ^2^ = 23.83, *p* < .001). 22/30 participants showed a meaningful increase in map volume, which represented a significant proportion (χ^2^ = 6.53, *p*= .01), while 36/44 showed a meaningful decrease in map volume, which also represented a significant proportion (χ^2^ = 17.81, p< .001).

**Figure 7.**
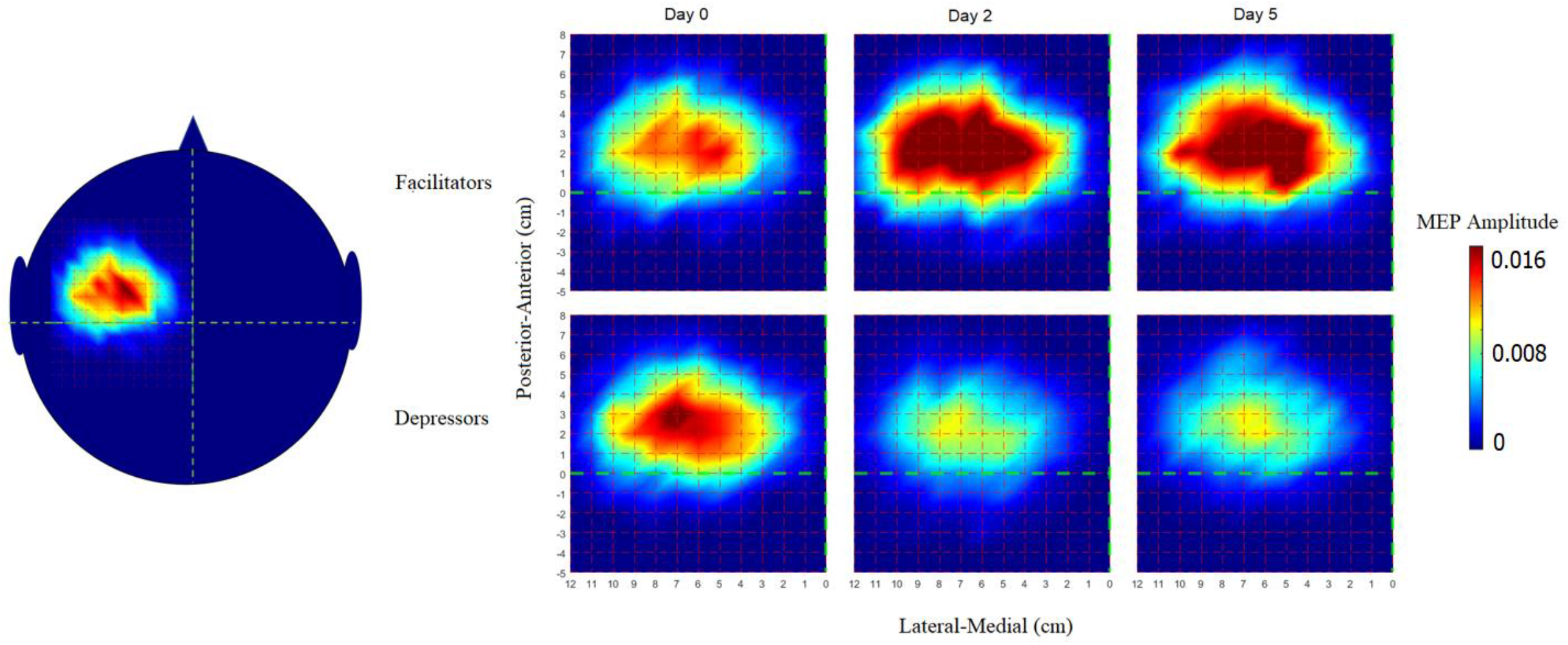
Motor cortex maps for facilitators (those who showed a mean increase in map volume across days, n = 30) and depressors (those who showed a mean decrease in map volume across days, n = 44). The map shows the mean MEP amplitude at each stimulation site relative to the vertex. The figure on the left shows roughly where TMS was applied over the scalp. The dashed green lines represent the horizontal and vertical lines over the vertex.

## Discussion

The aim of the present study was to determine a) whether PAF remains stable over several days of sustained pain, and b) whether individuals show two stable and distinct patterns of CME change during sustained pain. The EEG results showed moderate to excellent reliability in PAF, with excellent reliability observed when using a weighted frequency approach to calculate PAF. The TMS results supported the presence of two stable corticomotor responses to several days of NGF-induced muscle pain: corticomotor facilitation and depression.

### Reliability of PAF

Several studies have shown that PAF is highly stable over time [40-43]. The present study extends these findings by a) demonstrating that PAF remains stable even in the presence of experimentally induced pain and b) showing that the test-retest reliability of PAF during pain can be influenced by various methodological factors. The most critical factor was the peak identification method, with the centre of gravity method leading to excellent reliability estimates, and peak picking method leading to moderate to good reliability estimates in PAF. It is conceivable that the use of a single peak estimate is more prone to between sessions fluctuations given the potential presence of multiple peaks. Using a wider frequency window (8-12Hz) may introduce even more peaks within the calculation window and further inflate the fluctuations in PAF between sessions. In contrast, the use of a weighted peak estimate takes into account the presence of multiple peaks and is thus less susceptible to between-sessions fluctuations [44]. Interestingly, for both the centre of gravity and peak-picking methods, reliability was minimally affected by pre-processing pipeline or recording length. This is consistent with a recent study that showed similar PAF estimates between pre-processing pipelines [18], though this previous study did not investigate a “no pre-processing” pipeline that only involved re-referencing the signal, nor was between-sessions reliability during pain investigated. The lack of influence of pre-processing and recording length is likely due to the nature of the data collection method, wherein participants were instructed to relax their muscles with eyes closed, minimizing the influence of muscle and eye movement artefacts during the recording. As such, our results demonstrate that reliable PAF data can be obtained with minimal pre-processing and with recording lengths of ∼2 minutes. The relative ease at which PAF data can be obtained is promising for subsequent stages of biomarker development, including clinical validation and intervention studies.

### Feasibility of CME classification methods

The findings suggest that experimentally induced musculoskeletal pain sustained over several days is associated with two distinct and stable corticomotor responses: facilitation and depression. A large proportion of individuals showed a meaningful increase or decrease in corticomotor excitability, which was identified as a minimum of ∼17% change in map volume from Day 0. The percentage of the sample showing a mean increase (30/74 i.e. 40%) or decrease (44/74 i.e. 60%) in CME was similar to what was observed in a previous study using the NGF model in the elbow muscle [9]. As such, the results provide further support for the generalizability of distinct corticomotor responses to pain in different muscles with markedly different motor functions. It is important to note however, that no study has determined the proportion of participants showing *meaningful* increases or decreases in CME during pain, nor has any study determined whether these changes are stable. Many theories of chronic pain [21, 22] suggest that individuals show significant variations in motor strategies in response to pain, with some of these strategies associated with the transition to chronic pain. Our results provide novel characterisation of these individualised motor strategies to sustained pain. Further work is required to determine whether these strategies are involved in the transition to chronic pain.

### Limitations and Recommendations

One limitation of the present study was that recording length was manipulated offline. Further studies are required to determine whether reliable PAF data can be obtained at differing recording lengths manipulated online. These studies may also establish the shortest possible recording length for reliable PAF data. This could assist researchers and clinicians who wish to use efficient methods of classifying at-risk individuals using resting-state EEG. Further work is also required to investigate the reliability of PAF during eyes open, which is important for studies that assess PAF during task performance.

A further limitation of the study was the lack of assessment of CME beyond Day 5, especially given NGF-induced pain can last 2-3 weeks in some individuals [9]. Assessment of corticomotor excitability over a longer period may provide meaningful information on the stability of these corticomotor adaptations to pain, the amount of time that these adaptations last, and whether these adaptations persist after the resolution of pain.

## Conclusions

In summary, the present findings provide further support for the reliability of two prospective cortical pain biomarkers. Specifically, it was shown that PAF was reliable over three recording sessions, even in the presence of sustained pain, and even when considering various methodological factors that could influence PAF. Furthermore, it was shown that experimentally induced sustained pain produced two distinct corticomotor responses in the masseter muscle: facilitation and depression. This supports the goals of the ongoing PREDICT trial to validate these biomarkers as predictors of pain sensitivity.

## Data Availability

All data produced in the present study are available upon reasonable request to the authors

